## Supplementary Materials for "Pegylated-interferon-λ treatment-induced peripheral interferon stimulated genes are associated with SARS-CoV-2 viral load decline despite delayed T cell response in older individuals"

**Supplementary Table 1. scRNA sequencing patient characteristics**

| | Placebo | PEG-IFN-λ | Total |
|---|---|---|---|
| **Total #** | 4 | 5 | 9 |
| **Sex** | | | |
| Female | 2 | 3 | 5 |
| Male | 2 | 2 | 4 |
| **Median age, years (range)** | 47 (25-63) | 47 (27-60) | 47 (25-63) |
| **IFNL4 genotype** | | | |
| _G | 0 | 0 | 0 |
| TT/_G | 1 | 2 | 3 |
| TT | 3 | 3 | 6 |

**Supplementary Table 2. List of ISGs used in ISG score computation**

| ISG |
|---|
| IFIT1 |
| IFI6 |
| ISG15 |
| OAS1 |
| MX1 |
| OASL |
| STAT1 |
| JAK1 |
| IFI27 |
| IFITM1 |
| IFITM2 |
| IFITM3 |
| IFI44L |
| B2M |
| CD83 |
| DDIT4 |
| EHD4 |
| IFIT2 |
| IRF1 |
| NFKBIZ |
| NXPE3 |
| PIM3 |
| RSG2 |
| TNFSF10 |

**Supplementary Table 3. RBD-specific Ig correlations.**

| **Correlations (Spearman r)** | | | | | | | |
|---|---|---|---|---|---|---|---|
| **D7** | IgG | IgA | IgM | **D90+** | IgG | IgA | IgM |
| IgG | | | | IgG | | | |
| IgA | 0.4243* | | | IgA | 0.3226 | | |
| IgM | 0.7900**** | 0.6240*** | | IgM | 0.1083 | 0.1244 | |

* = correlation $p < 0.05$
*** = correlation $p < 0.001$
**** = correlation $p < 0.0001$

**Supplementary Table 4. RBD-specific IgG correlations with spike T cell responses**

| **Correlations (Spearman r)** | | | | | | | | | | | |
|---|---|---|---|---|---|---|---|---|---|---|---|
| **D0** | IgG | IgA | IgM | **D7** | IgG | IgA | IgM | **D90+** | IgG | IgA | IgM |
| IFN-γ | 0.3001 | 0.3875 | 0.1956 | IFN-γ | 0.2769 | 0.1119 | 0.1654 | IFN-γ | **0.4548*** | **0.5470**** | -0.1418 |
| IL-2 | 0.3364 | 0.2067 | 0.0568 | IL-2 | 0.1277 | 0.0269 | 0.0146 | IL-2 | **0.5559**** | **0.4950*** | -0.0041 |
| Poly | 0.1781 | 0.3267 | 0.1600 | Poly | 0.2623 | 0.0935 | 0.1562 | Poly | **0.5115*** | **0.5537**** | -0.0564 |

* = correlation $p < 0.05$
** = correlation $p < 0.01$

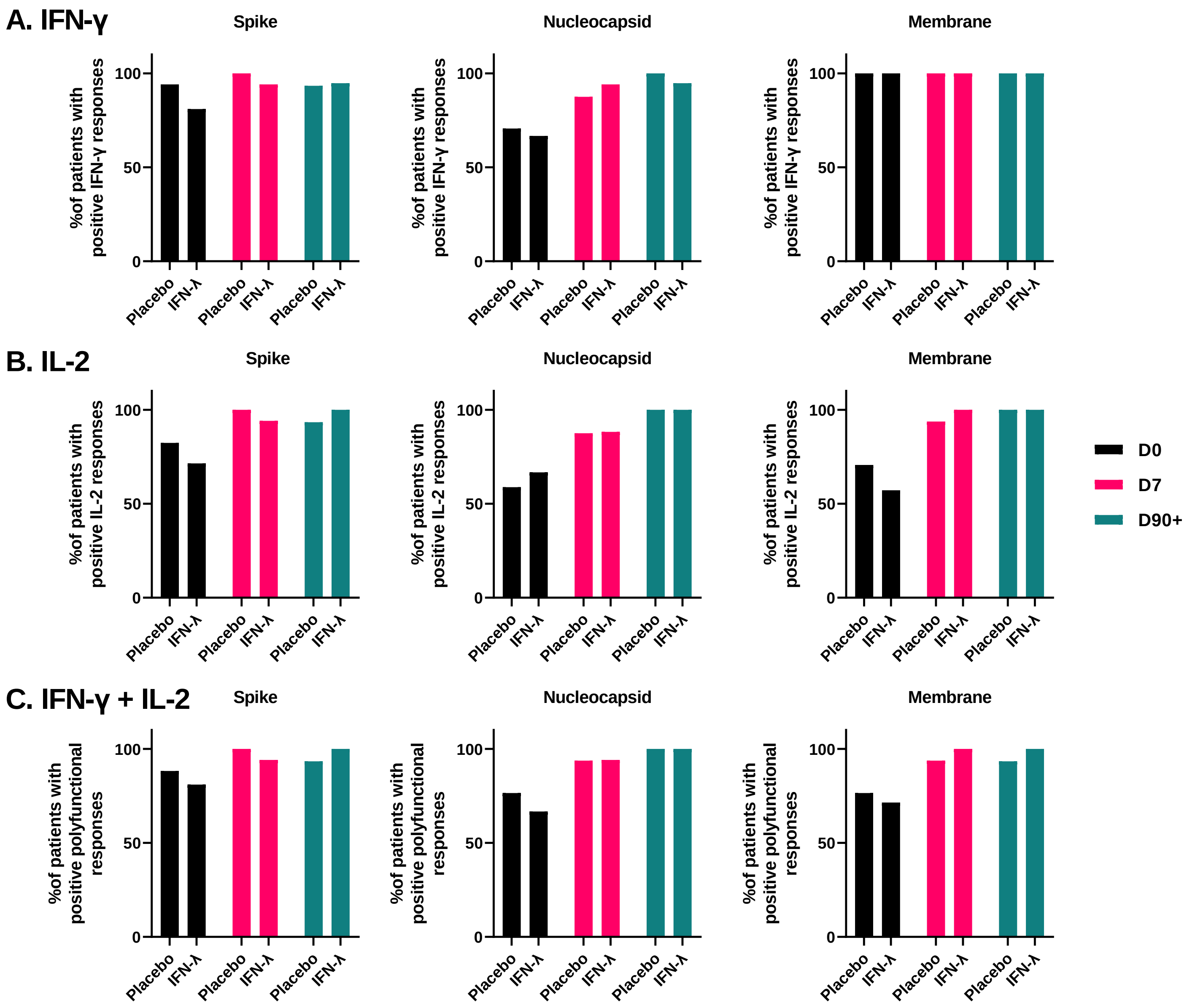


**Supplementary Figure 1. Percentage of positive A) IFN-γ B) IL-2 and C) polyfunctional T cell responses towards SARS-CoV-2 structural proteins across D0, D7, and D90+.** Responses were determined by fluorospot assay, with positive responses defined as the number of spot forming units (SFUs)/million exceeding twice the individual's negative control SFU count and greater than the mean negative SFU count from all patients. No significant difference in proportions between treatment groups was observed (Chi-square test with Yates' correction).

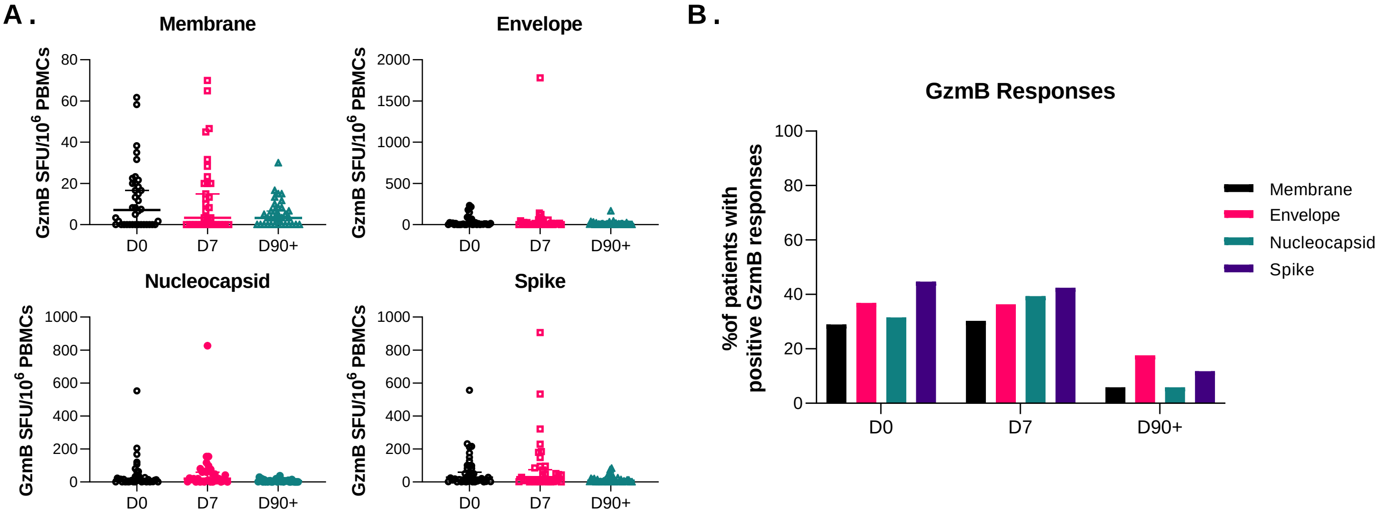


**Supplementary Figure 2. GzmB T cell responses to SARS-CoV-2 structural proteins.** A) The number of SFUs/$10^6$ PBMCs according to each SARS-CoV-2 protein B) The percentage of patients with a positive GzmB response towards the SARS-CoV-2 proteins. Each dot represents a different patient. No significant differences between groups (Mann-Whitney U tests). Bar lines represent median and 95% CI.

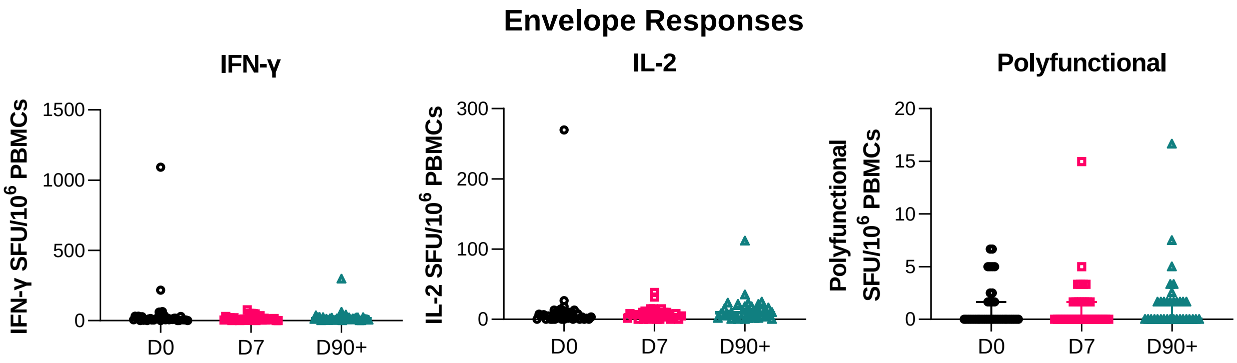


**Supplementary Figure 3. T cell responses to SARS-CoV-2 envelope protein.** IFN-γ, IL-2 and polyfunctional (IFN-γ + IL-2) T cell responses (as SFUs per $10^6$ PBMCs) against SARS-CoV-2 envelope peptide pools were quantified *ex vivo* using fluorospot assays. Each dot represents a different patient. Bar lines represent median and 95% CI.

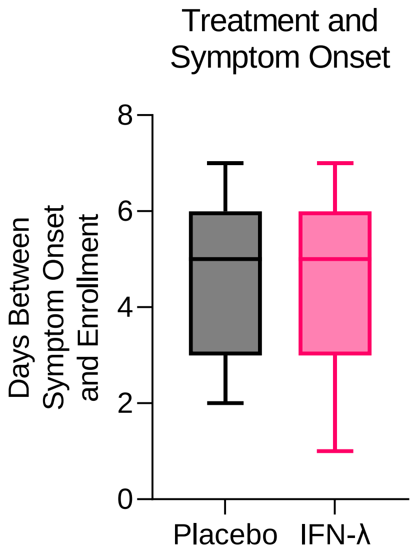


**Supplementary Figure 4. Differences in the time between symptom onset and enrollment in different treatment groups.** Bar lines represent median and 95% CI.

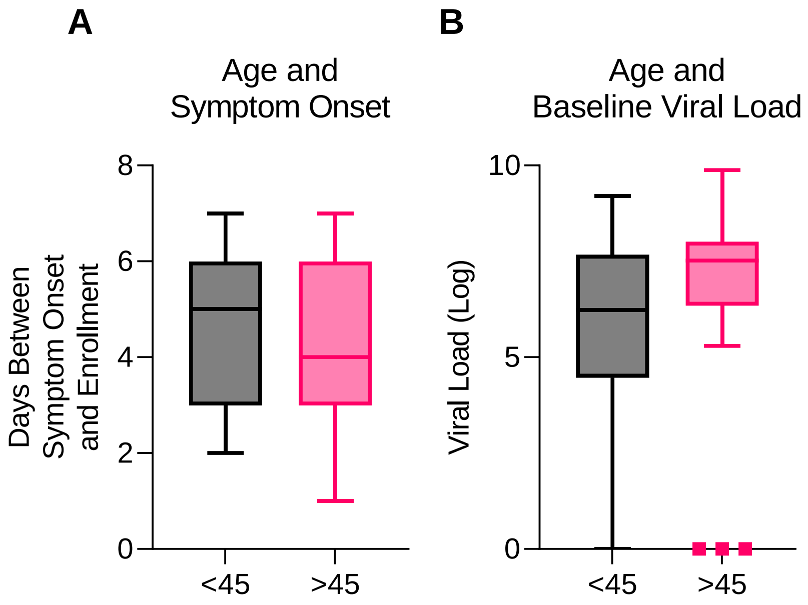


**Supplementary Figure 5. Differences in age and A) the time between symptom onset and enrollment and B) baseline viral load.** Bar lines represent median and 95% CI.

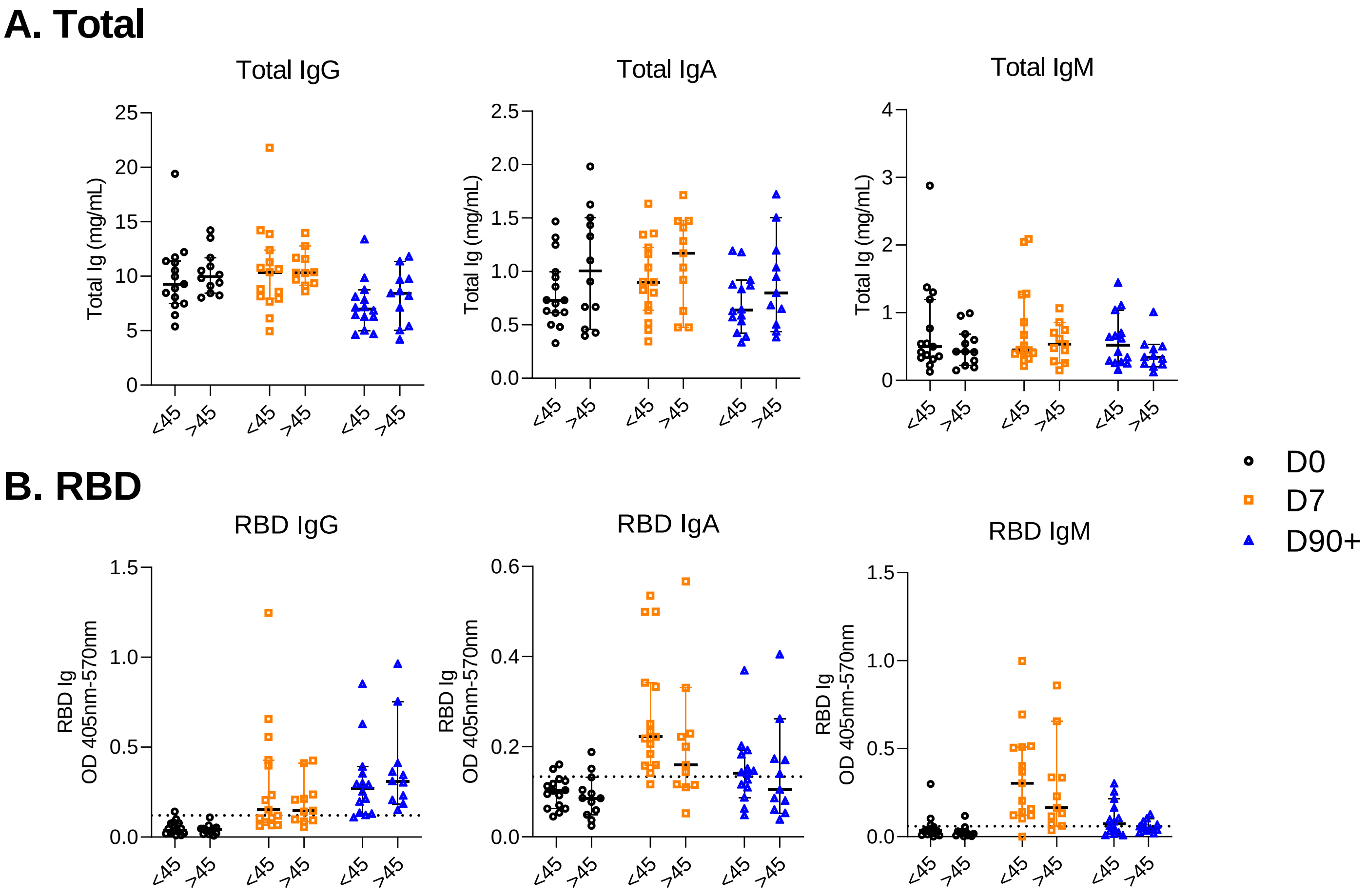


**Supplementary Figure 6. Differences in A) total and B) RBD-specific antibody between patients below and above 45 years old at day 0, day 7, and day 90+.** Dashed line in B) represents the mean + 2SD of results obtained from 8 pre-pandemic plasma controls collected in 2018-2019. Each dot represents a different patient. Bar lines represent median and 95% CI.

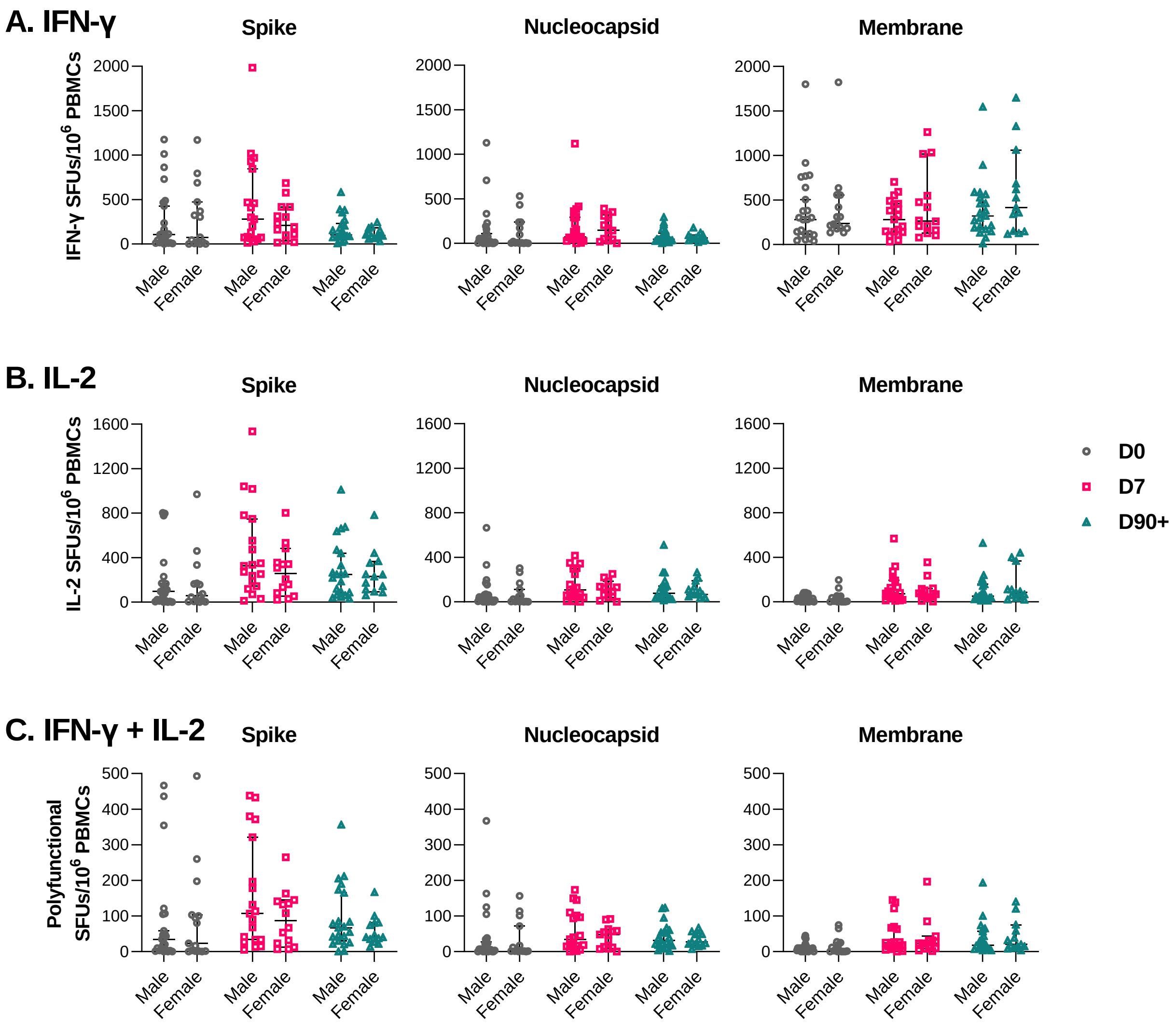


**Supplementary Figure 7. Differences in T cell responses between sex at day 0, day 7, and day 90+.** A) IFN-γ B) IL-2 C) Polyfunctional (IFN-γ + IL-2) T cell responses (as SFUs per $10^6$ PBMCs) against structural SARS-CoV-2 protein peptide pools were compared between male and females. Significant differences were not observed using Mann-Whitney U-tests between sexes ($p < 0.05$). Bar lines represent median and 95% CI.

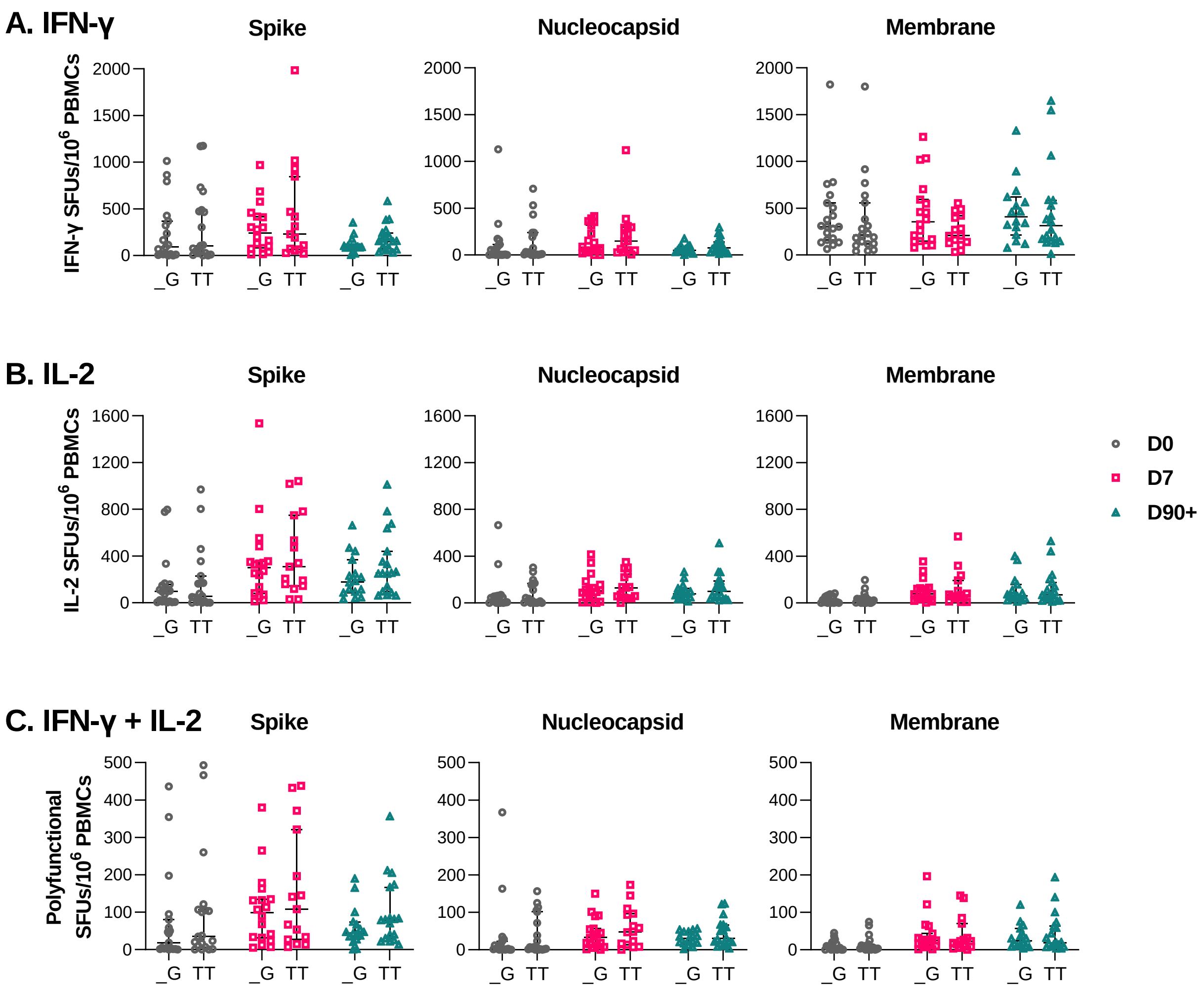


**Supplementary Figure 8. Comparison of T cell responses between *IFNL4* genotype at day 0, day 7, and day 90+ post-enrollment.** A) IFN-γ B) IL-2 C) Polyfunctional (IFN-γ + IL-2) T cell responses (as SFUs per $10^6$ PBMCs) against structural SARS-CoV-2 protein peptide pools were compared between genotypes. "_G" indicates non-TT rs368234815 polymorphisms at the IFNL4 locus. Significant differences were not observed using Mann-Whitney U-tests between genotypes ($p > 0.05$). Bar lines represent median and 95% CI.

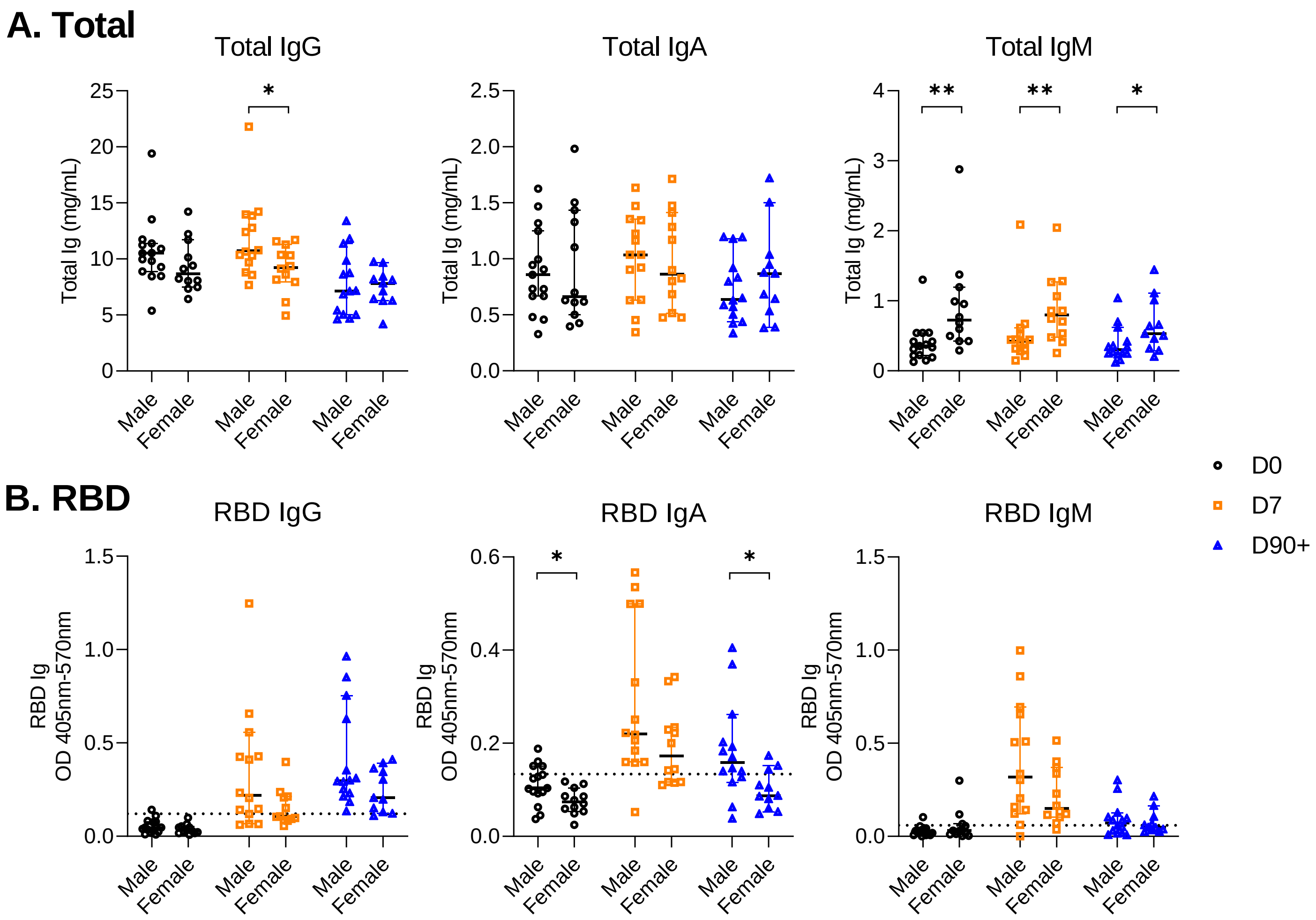


**Supplementary Figure 9. Differences in A) total and B) RBD-specific antibody levels between sex at day 0, day 7, and day 90+.** Dashed line in B) represents the mean + 2SD of results obtained from 8 pre-pandemic plasma controls collected in 2018-2019. Each dot represents a different patient. Significant differences were observed using Mann-Whitney U-tests between sexes (*p < 0.05). Bar lines represent median and 95% CI.

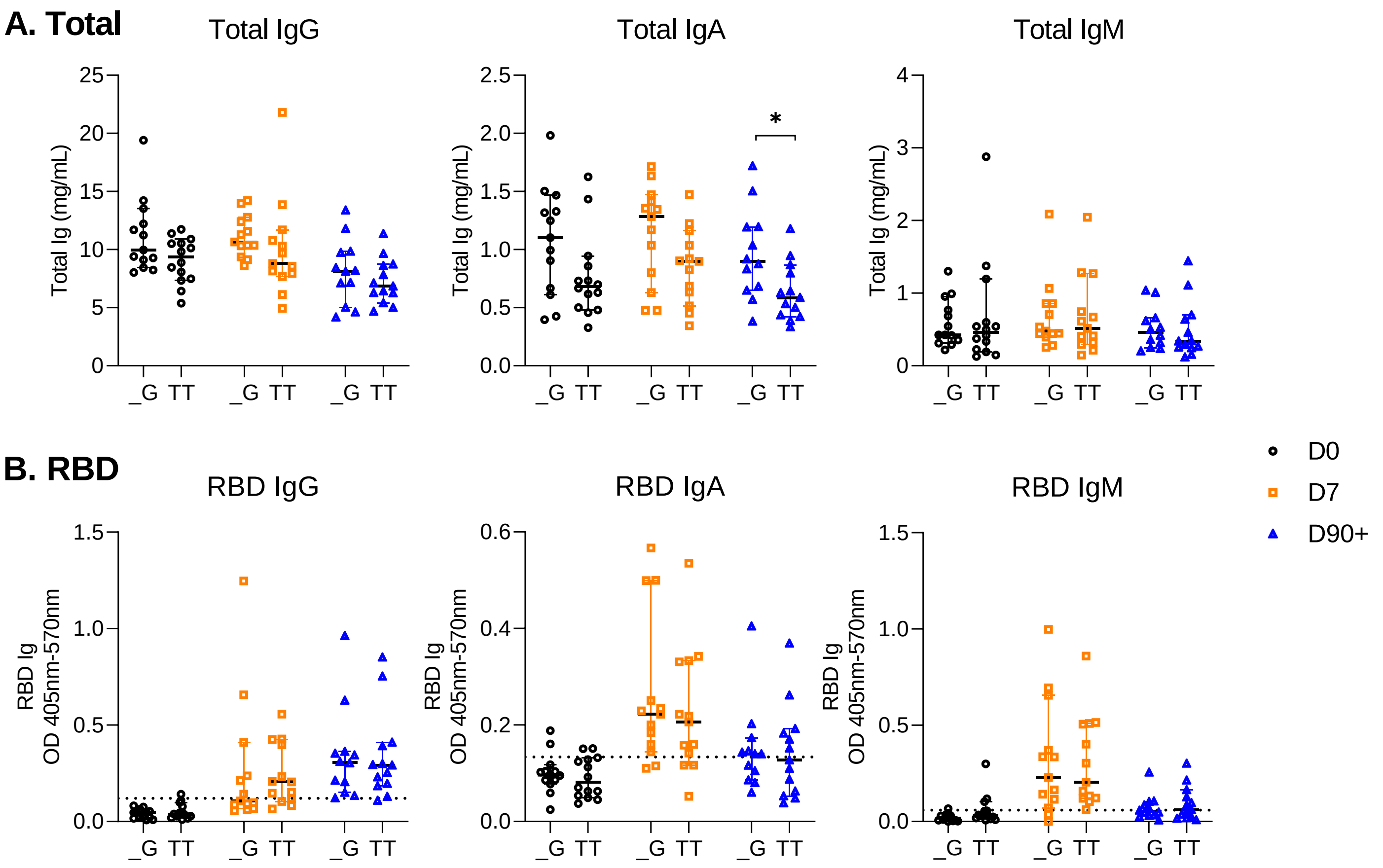


**Supplementary Figure 10. Differences in A) total and B) RBD-specific antibody levels between *IFNL4* genotype at day 0, day 7, and day 90+.** Significant differences were observed using Mann-Whitney U-tests between IFNL4 genotypes (* p < 0.05). Bar lines represent median and 95% CI.
